# Detection and quantification of SARS-CoV-2 RNA in wastewater influent in relation to reported COVID-19 incidence in Finland

**DOI:** 10.1101/2021.10.05.21264462

**Authors:** Ananda Tiwari, Anssi Lipponen, Anna-Maria Hokajärvi, Oskari Luomala, Anniina Sarekoski, Annastiina Rytkönen, Pamela Österlund, Haider Al-Hello, Aapo Juutinen, Ilkka T. Miettinen, Carita Savolainen-Kopra, Tarja Pitkänen

## Abstract

Wastewater-based surveillance is a cost-effective concept for monitoring COVID-19 pandemics at a population level. Here, SARS-CoV-2 RNA was monitored from a total of 693 wastewater (WW) influent samples from 28 wastewater treatment plants (WWTP, N = 21–42 samples per WWTP) in Finland from August 2020 to May 2021, covering WW of ca. 3.3 million inhabitants (∼ 60% of the Finnish population). The relative quantity of SARS-CoV-2 RNA fragments in the 24h-composite samples was determined by using the ultrafiltration method followed by nucleic acid extraction and RT-qPCR assay targeted with N2-assay. SARS-CoV-2 RNA signals at each WWTP were compared over time to the numbers of new and confirmed COVID-19 cases in the sewer network area.

Over the 10-month surveillance period, the detection rate of SARS-CoV-2 RNA in WW was 79% (including 6% uncertain results), while only 24% of all samples exhibited gene copy (GC) numbers above the quantification limit. The range of the SARS-CoV-2 detection rate in WW varied from 33% (including 10% uncertain results) in Pietarsaari to 100% in Espoo. Only six out of 693 WW samples were positive with SARS-COV-2 RNA when the reported COVID-19 case number from the preceding 14 days was zero. Overall, the 14-day COVID-19 incidence was 7, 18 and 36 cases within the sewer network area when the probability to detect SARS-CoV-2 RNA in wastewater samples was 50%, 75% and 95%, respectively. The quantification of SARS-CoV-2 GC required significantly more COVID-19 cases: the quantification rate was 50%, 75% and 95% when the 14-day incidence was 110, 152 and 223 COVID-19 cases, respectively, per 100 000 persons. Multiple linear regression confirmed the relationship between the COVID-19 incidence and the SARS-CoV-2 GC quantified in WW at 15 out of 28 WWTPs (overall R^2^ = 0.36, *p* < 0.001). At four of the 13 WWTPs where a significant relationship was not found, the GC of SARS-CoV-2 RNA remained below the quantification limit during the whole study period. In the five other WWTPs, the sewer coverage was less than 80% of the total population in the area and thus the COVID-19 cases may have been inhabitants from the areas not covered.

Based on the results obtained, WW-based surveillance of SARS-CoV-2 could be used as an indicator for local and national COVID-19 incidence trends. Importantly, the determination of SARS-CoV-2 RNA fragments from WW is a powerful and non-invasive public health surveillance measure, independent of possible changes in the clinical testing strategies or in the willingness of individuals to be tested for COVID-19.

## 1. Introduction

Wastewater-based epidemiology (WBE) has been used in areas with centralized sewage network systems for evaluating the circulation of etiological agents of communicable diseases such as hepatitis C virus, poliovirus, and antibiotic-resistant bacteria, consumption patterns of illegal drugs, nicotine, alcohol, and pharmaceuticals in communities (Gracia-Lor et al. 2017, Lorenzo and Picó 2019, Sims et al. 2020). Recently, WBE has been reported as a quick, sensitive, and cost-effective approach for monitoring the prevalence, trend, and circulation of coronavirus disease (COVID-19) pandemics at the population level (Medema et al. 2020a, Hart and Halden 2020, Ahmed et al. 2020, Sherchan et al. 2020, Wang et al. 2020, Gonzalez et al. 2020, Hillary et al. 2021, Rusiñol et al. 2021, Lundy et al. 2021), and as an early warning tool (Medema et al. 2020a, Wu et al. 2020a, Ahmed et al. 2021a).

During the COVID-19 pandemic in 2020–2021, the WBE approach has been used worldwide to complement the clinical (individual testing) surveillance approach (Ahmed et al. 2020, Kumar et al 2020, WHO 2020, Sherchan et al. 2020, Hokajärvi et al. 2021). Sometimes, WBE data can be more reliable than clinical data, as the clinical diagnostic capacity is limited mostly to the population having symptoms or with a recent travel history (Wu et al. 2020a). Further, clinical data can be biased depending on various factors such as differences in patient testing strategies, and sometimes the unwillingness of people to be tested. In that respect, the WBE approach is more unbiased as it accounts for the viral load of all infected (i.e., symptomatic, asymptomatic, pre-symptomatic, and post-symptomatic) people within the sewer network area (Wu et al. 2020b, Cevik et al. 2021, Wölfel et al. 2020).

The WBE approach to monitoring COVID-19 is an area of rapid development and thus all the factors defining the minimum threshold number of new COVID-19 cases within a sewer network area for detecting SARS-CoV-2 in the influent WW are not clear. One of the necessary considerations from the clinical standpoint is the variability in shedding quantities (viral load) and secretion routes (feces, urine, cough, sneeze, and sputum) of infected individuals from where virus particles end up in the sewage systems (Wölfel et al. 2020, Wang et al. 2020, Wu et al. 2020, Cevik et al. 2021). From the environmental standpoint, the fate and decay of SARS-CoV-2 in sewer networks and transit after sampling before analysis are not fully known (Hart and Halden 2020). Further, runoff waters and industrial WW might dilute SARS-CoV-2 quantities, and therefore various normalization procedures of SARS-CoV-2 copy numbers in WW are used; most often in the form of flow and population size normalization, but microbial indicators of human fecal loads have also been proposed (Medema et al. 2020a, 2020b, Green et al. 2020). Overall, a better understanding of the relationship between community COVID-19 incidence and SARS-CoV-2 GC in WW is needed for further development of the WBE approach.

Herein, this study compared a 10-month (August 2020 to May 2021) longitudinal monitoring of SARS-CoV-2 RNA in WW influent samples analyzed from 28 WWTPs in Finland with a 14-day COVID-19 case incidence rate (14-day moving sum, 14-DMS) of new and confirmed COVID-19 cases in the respective communities. The minimum number of COVID-19 cases needed in the respective communities for detecting and quantifying SARS-CoV-2 RNA in WW influent samples in Finland was determined and the potential of WBE to catch the local and national COVID-19 incidence trends was investigated.

## 2. Materials and Methods

### 2.1 Wastewater sample collection

Between 3 August 2020 and 31 May 2021, a total of 693 influent WW samples were collected following the standard biosafety precautions for handling untreated WW as previously described (Hokajärvi et al. 2021). Samples were collected from 28 wastewater treatment plants (WWTPs) serving about 3.3 million inhabitants; this is about 60% of the total population of Finland.

Population coverage between the participating WWTPs varied from 860 000 inhabitants in Helsinki to 18 000 in Vihti, causing variability between WWTPs in the mean influent flow during the sampling events (Table 1). The mean WW influent inflow normalized per 100 000 inhabitants was 29 000 m^3^/day during the 24-hour composite sampling events. A fraction (∼ 1 liter) of samples were transported in cool boxes as soon as possible to the Water Microbiology Laboratory of the Finnish Institute for Health and Welfare (THL), Kuopio, Finland, for analysis. As soon as the sample arrived at the laboratory, the arrival time and temperature were recorded, and the samples were stored at + 4 °C and mostly analyzed within 24–48 hours.

**Table 1.** Wastewater influent samples collected at the 28 WWTPs between 3 August 2020 and 31 May 2021, Finland.

| WWTP, location | N | 24h-composite sampling |  | Industrial WW | 24-h WW influent volume | Temperature, Median (Min-Max) |  | Transit time (days) | TSS (mg/l) |
| --- | --- | --- | --- | --- | --- | --- | --- | --- | --- |
| | | Method | Interval | Proportion, % | Mean ± SE, * | At WWTP, °C | After transit, °C | Median (Min-Max) | Mean (Min-Max), N |
| Viikinmäki, Helsinki | 42 | Flow/time | 3x/10000 m<sup>3</sup>/ 20 min | 7 | 34 160±1 000 | 13.7 (9.7-18.5) | 10.8 (7.4-18.3) | 1 (1-2) | 298.6 (128-738), 42 |
| Suomenoja, Espoo | 22 | Flow | 750 m<sup>3</sup> | 5 | 27 760±1 190 | 13.4 (8.5-18.1) | 8.9 (5.7- 14.5) | 1 (1-2) | 295.5 (220-416), 22 |
| Kakolanmäki, Turku | 42 | Flow | 990 (700-1500) m<sup>3</sup> | 15 | 26 780±1 530 | 12.1 (8.4-20.1) | 10.7 (6.3- 21.4) | 1 (1-2) | 293.6 (176-448), 20 |
| Taskila, Oulu | 41 | Time | 38 min | NA | 25 880±770 | 9.1 (6.7-15.7) | 10.4 (5.6- 22.0) | 1 (1-5) | 457.9 (230-690), 19 |
| Viinikanlahti, Tampere | 41 | Flow | NA | NA | 33 320±850 | 16.2 (11.6-23.5) | 10.2 (3.7- 20.9) | 1 (1-6) | 437.8(220-970), 41 |
| Nenäinniemi, Jyväskylä | 21 | Flow | 280 (150-550) m<sup>3</sup> | 10 | 23 610±1 110 | 11.5 (7-18.2) | 11.2 (6.6- 18.4) | 1 (1-2) | 740 (380-1080), 21 |
| Luotsinmäki, Pori | 21 | Flow | 250 m<sup>3</sup> | 10 | 26 880±860 | 10.4 (7.9-17.4) | 11.1 (6.6- 20.8) | 2 (1-2) | NA |
| Mussalo, Kotka | 21 | Flow | NA | 13 | 30 560±2 080 | 12.4 (7.4-19.1) | 9.9 (5.6- 21.6) | 2 (1-2) | 390 (390-390), 1 |
| Kuhasalo, Joensuu | 21 | Flow | 110 (100-200) m<sup>3</sup> | 20 | 20 490±920 | 10.4 (6.9-16) | 10.6 (5.7- 16.4) | 1 (1-2) | 280.7 (240-400), 14 |
| Lehtoniemi, Kuopio | 42 | Flow/time | 1 100 m<sup>3</sup>/ 60 min | NA | 24 540±730 | 11.1 (7.5-16.8) | 10.4 (6.0- 17.1) | 1 (0-3) | 460 (420-500), 2 |
| Pätt, Vaasa | 22 | Flow | 2 000 m<sup>3</sup> | NA | 27 260±1 450 | 11.6 (7.6-18.4) | 9.0 (6.0- 16.7) | 1.5 (1-2) | NA |
| Mäkikylä, Kouvola | 22 | Flow | NA | 4.4 | 37 280±2 940 | 10 (5.9-15.3) | 9.6 (4.0- 20.6) | 1 (1-3) | 304.7 (206-520), 12 |
| Paroinen, Hämeenlinna | 21 | Flow | 540 (500-800) m<sup>3</sup> | NA | 25 470±1 170 | 9.5 (6.3-15.5) | 11.0 (7.0- 19.0) | 1 (1-2) | NA |
| Kariniemi, Lahti (I) | 21 | Flow | 180 m<sup>3</sup> | 15 | 27 670±1 320 | 13.5 (8-17.7) | 10.4 (6.4- 18.7) | 1 (1-2) | 289.2 (150-470), 12 |
| Toikansuo, Lappeenranta | 21 | Time | 10 min | 10 | 24 290±1 100 | 10 (4-16.2) | 9.4 (7.0- 19.3) | 1 (1-3) | 454.7 (200-1400), 21 |
| Ali-Juhakkala, Lahti (II) | 21 | Flow | 150 m<sup>3</sup> | 12 | 23 210±1 230 | 13.1 (7.7-17.4) | 10.8 (6.0- 18.7) | 1 (1-2) | 430 (220-740), 14 |
| Alakorkalo,<br>Rovaniemi | 21 | Flow | 1 m<sup>3</sup> | NA | 32 150±1 260 | 9.1 (6.8-14.4) | 11.7 (6.4- 21.1) | 1 (1-3) | NA |
| Keskuspuhdistamo,<br>Salo | 21 | Flow | 60 m<sup>3</sup> | NA | 41 330±13 080 | 9.2 (4.8-15.3) | 9.7 (6.5- 18.9) | 1 (1-15) | NA |
| Keskuspuhdistamo,<br>Seinäjoki | 21 | Flow | 400 m<sup>3</sup> | NA | 38 270±2 450 | 10 (7-16) | 10.4 (5.4- 18.6) | 2 (1-3) | 171.2 (96-310), 5 |
| Kenkäveronniemi,<br>Mikkeli | 21 | Flow | 50 m<sup>3</sup> | NA | 24 010±1 590 | 11.6 (8.6-17.8) | 10.8 (6.7- 17.4) | 1 (1-3) | 540.8 (440-670), 13 |
| Maanpäänniemi,<br>Rauma | 21 | Flow | 340 (250-500)<br>m<sup>3</sup> | 20 | 28 250±1 010 | 7.5 (4-15.5) | 10.0 (5.0- 17.9) | 1 (1-2) | NA |
| Hopeakivenlahti,<br>Kokkola | 21 | Time | 15 min | 0 | 28 100±1 650 | 9.3 (5.5-16) | 9.6 (5.6- 20.1) | 2 (1-2) | 189.7 (96-390), 13 |
| Peuraniemi,<br>Kajaani | 21 | Time | 80 min | 1.5 | 32 050±1 980 | 10.5 (4.8-16.6) | 13.0 (7.6- 20.6) | 2 (1-2) | 436.7 (240-670), 12 |
| Alheda,<br>Pietarsaari | 22 | Time | 15 min | NA | 32 390±1 840 | 8.6 (6-16.3) | 10.0 (4.7- 15.0) | 2 (1-8) | 320 (320-320), 1 |
| Peurasaari,<br>Kemi | 21 | Flow/time | 50 ml / 10 min | NA | 43 660±3 660 | 6.8 (4.8-16.6) | 10.4 (7.4- 17.7) | 2 (1-2) | 158.3 (49-270) 6 |
| Pihlajaniemi,<br>Savonlinna | 21 | Time | 30 (20-40)<br>min | NA | 33 910±2 170 | 8.2 (4.1-17.8) | 12.0 (7.7-19.3) | 1 (1-3) | 197.9 (80-310), 21 |
| Lotsbroverket,<br>Maarianhamina | 21 | Flow | 50 m<sup>3</sup> | 30 | 27 870±2 250 | 9.9 (7-17.5) | 11.2 (7.2-20.1) | 2 (1-3) | NA |
| Nummela,<br>Vihti | 21 | Flow | 20 m<sup>3</sup> | NA | 17 600±470 | 11.7 (8-16.3) | 10.2 (7.3-21.4) | 2 (1-2) | 486 (390-580), 5 |
| **Total** | **693** |  |  | **10.3 (0-30)** | **29 230±540** | **11.3 (3.5-23.5)** | **10.4 (3.7- 22.0)** | **1 (0-15)** | **373.8 (49-1400), 317** |
| *m<sup>3</sup>/100 000 population; measured during the sampling events. Figure S3 shows the wastewater inflow volumes of the WWTPs without and with population normalization. N; number of samples. TSS; total suspended solids. NA; not available. |  |  |  |  |  |  |  |  |  |

The mean WW temperature was higher than 15 °C after transit to the laboratory in August 2020 and a part of the samples also exceeded this temperature limit in September and October 2020. The arrival temperature was mostly below 15 °C from November 2020 to May 2021. The time in transit varied between one and two days depending on the location (Table 1). The time in transit was longer than two days only for nine out of 693 samples.

### 2.2 Recording of the new COVID-19 cases in the WWTPs sewer network areas

Throughout the study period, individual COVID-19 tests have been available for all symptomatic people in Finland. Clinical laboratories report all detected COVID-19 cases to the National Infectious Disease Register (NIDR) detailing the total number of COVID-19 tests performed each day per each hospital district in Finland. The trend of daily reported cases of COVID-19 in the study sewer network areas during the study period shows increasing and decreasing trends (Figure 1).

**Figure 1.**
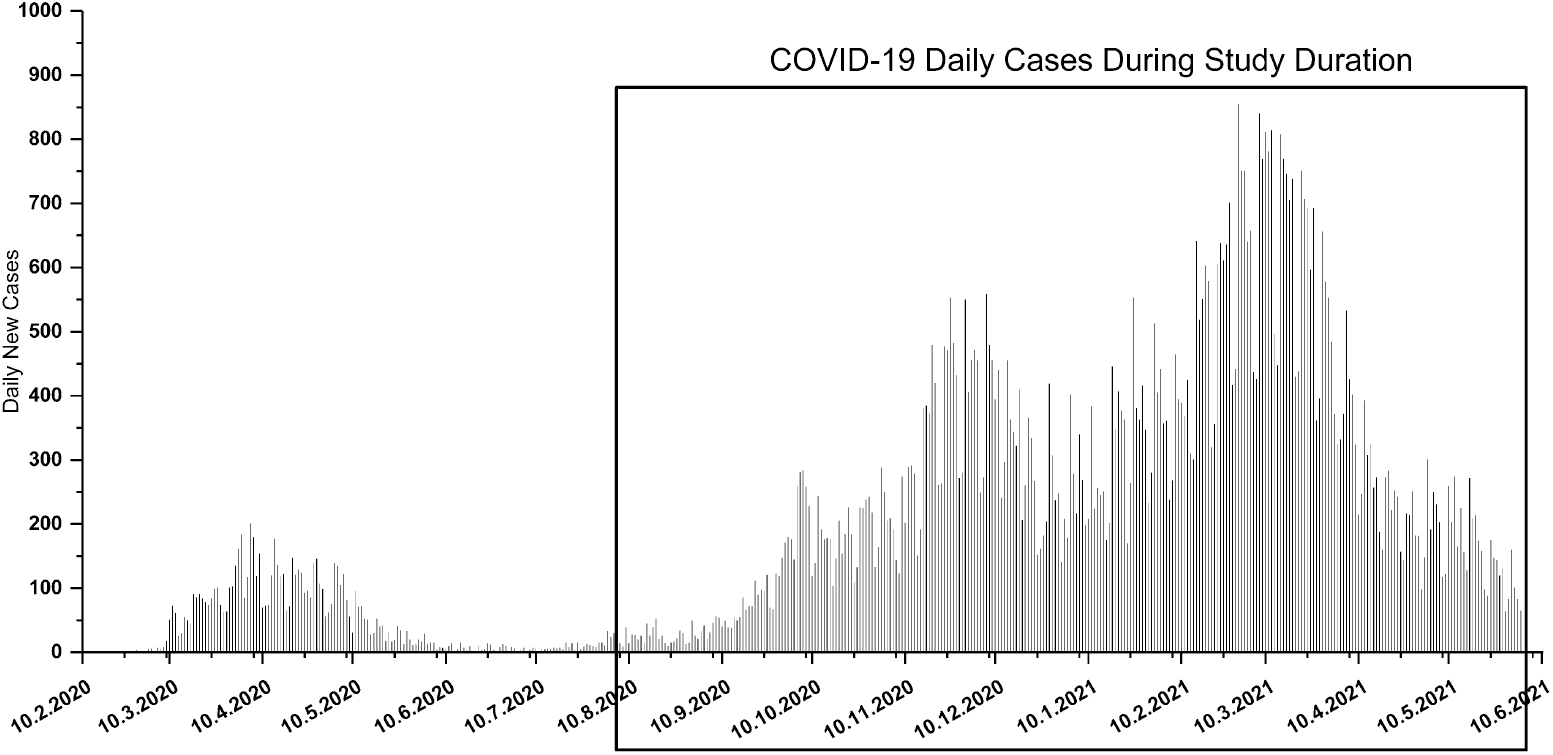
Total daily reported COVID-19 cases in the municipalities covered in the national wastewater sample collection.

For the purposes of comparing the clinical findings with the data produced by environmental monitoring of influent WW, the reported new COVID-19 cases in each municipality served by the 28 WWTPs were extracted from NIDR. Then the case numbers were corrected using a WWTP-specific factor corresponding to the share of inhabitants served by the sewer connected to the study WWTPs as compared to all inhabitants of the municipalities. This correction of the NIDR case numbers was necessary since each WWTP served one or more municipalities, and one municipality could have one or more WWTPs. Finally, the 14-DMS of the COVID-19 cases was calculated for each sewer network area. To compare with the WW data, the 14-day time window’s end was set to the day of composite sampling. These moving sum COVID-19 case numbers were then normalized per 100 000 population and we refer to these numbers as 14-day COVID-19 case incidence rate and in short 14-DMS.

### 2.3 Determination of SARS-CoV-2 RNA fragments in the wastewater samples

WW samples were analyzed as previously described (Hokajärvi et al. 2021). In brief, the ultrafiltration method (Medema et al., 2020) was used with the exception that 10 kDa Centricon Plus-70 centrifugal filters were used for 70 ml pre-centrifugated supernatants with a concentration-time of 25 minutes in 3 000 g producing 200 µl – 1 600 µl of concentrate. Mengovirus and crAssphage were used as internal process controls (Pintó et al. 2009, Stachler et al. 2017). Sterile deionized water was used as negative process control.

For nucleic acid extraction from 300 μl of the concentrate, and from 300 µl WW without ultrafiltration, a Chemagic Viral300 DNA/RNA extraction kit was used with the Chemagic-360D instrument (Perkin-Elmer, Germany). To verify the extraction performance, each extraction set included a positive swab sample (300 µl of 1:500 diluted, Ct approximately 29 after dilution, nasopharyngeal swab from a COVID-19 positive patient, dissolved into PBS and inactivated at 60°C for 90 min) and a negative extraction control (300 µl sterile deionized water).

All RT-qPCR and qPCR assays were performed using QuantStudio 6 Flex real-time PCR system (Applied Biosystems, ThermoFisher Scientific). In addition to the negative ultrafiltration and nucleic acid extraction process controls, all runs included at least one reaction with molecular-grade water instead of nucleic acid (no template control, NTC). RT-qPCR assay targeting nucleocapsid (N) protein gene of SARS-CoV-2 was used (N2 assay; Lu et al. 2020). For samples taken between August 2020 and the end of January 2021, also a beta-coronavirus assay to detect the envelope (E) protein gene was used (E-Sarbeco assay; Corman et al. 2020). The reactions and the target quantification were carried out as described earlier (Hokajärvi et al. 2021) by using TaqMan Fast Virus 1-step Master Mix (Applied Biosystems, ThermoFisher Scientific). Non-diluted and 10-fold diluted fractions of the extracted nucleic acid of each WW sample were analyzed in duplicates.

Mengovirus internal process control results to estimate the recovery efficiency and RT-qPCR inhibition were produced following the principles of international standard ISO/TS 15216-1:2013 (Pintó et al. 2009; Tables S2–S4). To characterize the fecal content of WW samples and further evaluation of the recovery efficiency of ultrafiltration, the cross-assembly phage (crAssphage) copy numbers were enumerated before and after ultrafiltration using a qPCR assay (Stachler et al. 2017). The total reaction volume of 25 μl in crAssphage assay contained 5 μl of nucleic acid template, primers in a final concentration of 1 µM, probe at concentration of 0.08 µM, and 12.5 µl of Environmental Master Mix 2.0 (Applied Biosystems, ThermoFisher Scientific). Single, 10- and 100-fold dilutions of nucleic acid templates were used for qPCR analysis. The quantification of crAssphage was performed using a synthetic gene fragment containing primer annealing sites (Integrated DNA Technologies, Belgium) with eight standard points10^6^-10^0^ GC/µl.

By using QuantiStudio Real-Time PCR System-software (Applied Biosystems, ThermoFisher Scientific), the reaction was considered successfully amplified when the Ct value was below 40 with a threshold in N2 0.1, E-Sarbeco 0.2, mengovirus 0.04, and crAssphage 0.05. The SARS-CoV-2 results (N2 assay) were interpreted using four categories, as follows:

- **Non-detected**: when all out of four reactions (two undiluted and two 10-fold diluted nucleic acids as a template) did not have any amplification (i.e., Ct > 40).
- **Uncertain:** One out of four reactions had amplification with Ct < 40 but was not confirmed in repeated RT-qPCR analysis.
- **Detected:** when more than one RT-qPCR reaction was positive in the N2 assay but copy numbers were below the limit of quantification (LOQ; 50 GC per reaction).
- **Detected and quantified:** copy number of SARS-CoV-2 target per 100 ml WW sample was calculated when the target was detected and the copy number exceeded the LOQ.

E-Sarbeco assay results were interpreted in the same categories as N2-assay, except copy numbers were not calculated. CrAssphage results were composed of arithmetic mean values of the two dilutions and reported as gene copy numbers per 100 ml of WW sample.

The presence of inhibition was reported as a factor decreasing the reliability of the result if the difference in Ct values was more than two in the mengovirus assay between the nucleic acid templates from a sample and a negative process control. In 63% of the samples tested (433 samples out of 684) the presence of inhibitors was noted. As the inhibition was prevalent in WW nucleic acids based on the mengovirus control, no samples were excluded from the data due to the inhibition. However, for the N2 assay both un-diluted and 10-fold diluted and for crAssphage 10- and 100-fold diluted nucleic acids were used to overcome the potential inhibitory effects in generating the results.

### 2.4 Cell cultures to determine the viability of SARS-CoV-2 in the wastewater samples

To determine the viability of SARS-CoV-2 in the WW samples using Vero E6 cell cultures, one WW influent sample collected at 10–11 May 2020 from WWTP in Helsinki and five WW influent samples collected at 18–19 October 2020 from WWTPs in Helsinki, Espoo, Vaasa, Jyväskylä, and Kouvola were used. Centricon concentrates were stored at −20 °C with penicillin-streptomycin-gentamycin antibiotics (final concentration100 IU/ml, 500 µg/ml each) prior to analysis.

A Centricon concentrate (3.0 ml, equivalent to 84 ml sample volume) obtained from the May 2020 sample was used (SARS-CoV-2 RNA copy number 11 000 GC/100 ml, N2 assay). Vero E6 cell lines were cultivated in minimum essential medium (MEM) with 2% fetal bovine serum (FBS), penicillin and streptomycin, and were incubated at 37 °C with 5% CO2 for 7 days. The concentrate was inoculated in five Vero E6 cell culture flasks (0.5 ml concentrate /4.5 ml MEM/flask). After the seventh day, the supernatant was filtered, and aliquots of 1.0 ml cell culture supernatant was inoculated on two fresh cell flasks for the second passage as described above. To determine the viability of the virus, each day the flasks were examined under the light microscope and 250 µl supernatant was collected for RNA extraction.

The SARS-CoV-2 RNA copy numbers of the October 2020 concentrates (N2 assay) were ca. 3 900 GC/100 ml (Helsinki), 6 600 GC/ 100 ml (Espoo), 20 000 GC/100 ml (Vaasa), 35 000 GC/100 ml (Jyväskylä) and 6 200 GC/100 ml (Kouvola). Concentrate volumes of 0.4 ml (equivalent to 40 ml sample volume) were used for cell cultures. The samples were filtrated (0.22 µm pore size) before inoculated on Vero E6 cells growing in 25 cm^2^ cell culture flasks for 1h at 37°C and 5 ml of fresh culture medium Eagle minimal essential medium (Eagle-MEM) (Sigma-Aldrich) supplemented with 2% fetal bovine serum (Sigma-Aldrich) was added for incubation. The cytopathic effect was observed under a light microscope daily during the six days of culture. After that, as a second passage, an aliquot of each cell culture supernatant was inoculated on fresh cells the same way as described earlier. A volume of 100 μl of the supernatant samples was collected daily for RNA extraction.

In addition, one WW sample was collected on 12 April 2021 during the peak of the epidemic from Helsinki WWTP and was used fresh to test the virus infectivity in the Vero E6 cell culture. A 500 ml WW sample was concentrated with dextran-polyethylene glycol (PEG)-mixture following the standard procedure described in the Polio Laboratory Manual (Hovi et al., 2001). Both the original and concentrated samples were further filtrated using 0.22 µm pore size membrane filters and incubated with penicillin-streptomycin-gentamycin antibiotics for 15 min at room temperature. Aliquots of both samples (100 µl of the concentrate equivalent to 3.3 ml of the original WW sample) were diluted in 1:1 with the culture medium supplemented with 2% fetal bovine serum (Sigma-Aldrich) and inoculated in five replicates on the Vero E6 cells growing in 2 cm^2^ cell culture wells (24-well plate) for 1h at 37°C after which 500 µl of fresh media was added on the cells. The cytopathic effect was observed under a light microscope daily during the three days of culture. After the third day, as a second passage, an aliquot of each cell culture supernatant was inoculated on fresh cells the same way as described earlier. A Finnish SARS-CoV-2 isolate hCoV-19/Finland/3/2020 (Gisaid number EPI_ISL_2365908) was used as a positive control. A volume of 100 µl aliquot of the supernatant samples was collected for RNA extraction on Day 0 and Day 3 from both passages.

The detection of SARS-CoV-2 RNA in the cell culture aliquots was done as described earlier (Jiang et al. 2021). Briefly, the RNA extraction was done with RNEasy mini kit (Qiagen), and detection of SARS-CoV-2 was done by using RT-qPCR assay targeting the E gene (Corman et al., 2020).

### 2.5 Data analysis and reporting of the SARS-CoV-2 results

Throughout the WW-based SARS-CoV-2 RNA monitoring efforts, the outcomes of the wastewater analysis were shared with governmental and local health authorities in Finland. The WW-based SARS-CoV-2 results were manually compared to reported new COVID-19 cases within the WWTP sewer network area municipalities. In case of any discrepancies between the clinical (individual testing) and environmental (wastewater testing) surveillance, direct contact by phone or email was made by THL’s personnel to the communicable disease doctor in charge of the corresponding hospital district. SARS-CoV-2 copy numbers of N2 assay were normalized with the influent flow at the sampled WWTP over each 24-h composite sampling event. Further, the numbers of inhabitants in the WWTP area were taken into consideration in presenting the SARS-CoV-2 RNA copy numbers, published weekly on THL’s website: (https://www.thl.fi/episeuranta/jatevesi/jatevesiseuranta_viikkoraportti.html).

To compare flowrate- and crAssphage-based normalization methods for N2 assay copy numbers, normalized copy numbers were calculated by dividing the N2 copy number per 100 ml by the crAssphage copy number per 100 ml of wastewater from all WWTP’s samples during the period from 1 November 2020 to 31 May 2021.

### 2.6 Statistical analysis

Statistical analysis was conducted using IBM SPSS statistics 27 and R (R Core Team, 2019). Figure illustrations were made using OriginPro (2017). Statistical tests were considered statistically significant when the p-value was < 0.05. For comparing SARS-CoV-2 RNA in WW influent to the 14-day COVID-19 case incidence rate, the 693 samples in total were converted into categorical interval data as follows: (a) at first, all the data tables were re-arranged based on ascending order of COVID-19 incidence, (b) then all samples with zero incidence cases were grouped into one category, (c) then categorical groups with 20 incidence case from lower to higher were categorized per group, and (d) out of 20 samples, the detection percentage of SARS-CoV-2 RNA per each interval was calculated; uncertain results were grouped into a detected category. For calculating the 14-day COVID-19 case incidence rate threshold values for detecting and quantifying SARS-CoV-2 RNA with N2 assay in WW influent, binary logistic regression analysis was used employing Hosmer-Lemeshow goodness of fit, where classification cutoff was 0.5 and maximum iterations 20.

By using the quantitative SARS-CoV-2 results in WW, linear regression analysis with multiple explaining variables for 14-day COVID-19 case incidence rate was conducted. In the preliminary data analysis for determining the most significant factor, COVID-19 incidence in the sewer network area, the number of customers per WWTP, wastewater influent inflow volume, sample collection month, and the sample temperature after transport all had a strong positive correlation (data not shown). Therefore, by avoiding the multicollinearity effect, we included only flow-corrected SARS-CoV-2 GC/day/100 000 persons, the temperature of the sample at arrival in the laboratory, and the number of days delay during sample transportation as explaining variables in the multiple linear regression analysis. In models for individual WWTPs, groups of large and small WWTPs, and all samples from 28 WWTPs pooled together, 14-day COVID-19 case incidence rate was used as the dependent variable. To include uncertain and detected results below LOQ to models, 25% of LOQ copy numbers for uncertain results and 50% for detected results were given and then normalized as described above. Flow-corrected copy numbers were replaced to linear models described above with copy numbers normalized against the sample crAssphage content to test crAssphage GC as a normalization method.

The Kruskal-Wallis test was used for determining the effect of population size of sewer network areas on SARS-CoV-2 detection in WW and 14-day COVID-19 case incidence rate, and sample transportation delays on the SARS-CoV-2 detection frequency in the influent wastewaters of WWTPs. The detection frequency between N2 and E-Sarbeco assays was compared with cross-tabulation followed by a Chi-square test by pooling samples from all WWTPs.

## 3. Results

### 3.1 National detection and quantification thresholds

The relationship between the 14-day COVID-19 case incidence rate and the WW-based SARS-CoV-2 detection and quantification rates was examined at the national level in Finland. COVID-19 incidence in the sewer network areas gradually increased the SARS-CoV-2 detection rate until reaching 100% in WW influent samples (Table 2, Figure 2). Based on the logistic regression analysis, the 14-day COVID-19 incidence was 7, 18 and 36 cases within the sewer network area when the probability to detect SARS-CoV-2 RNA in wastewater samples was 50%, 75% and 95%, respectively (Figure 3).

**Table 2.**
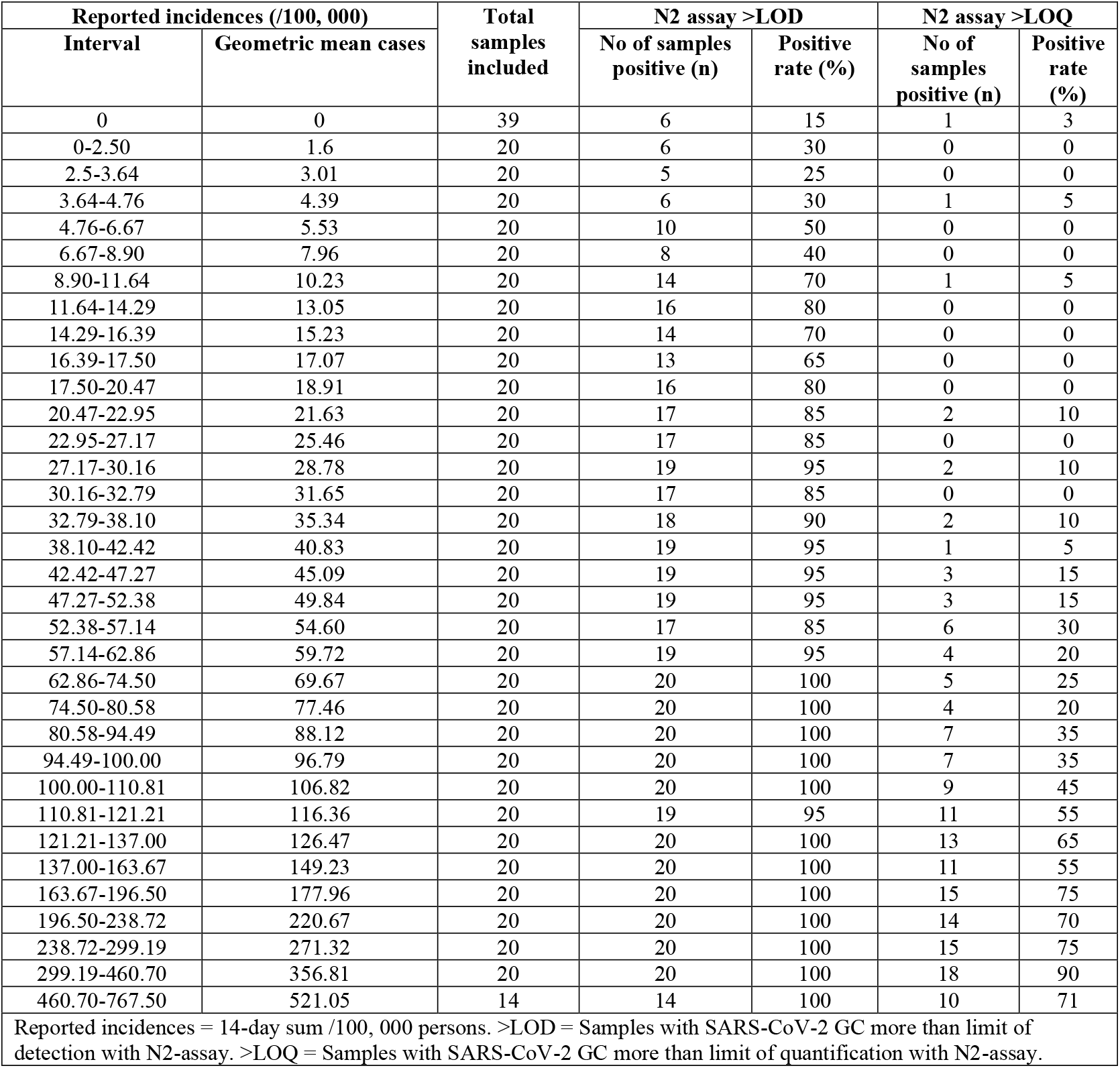
Detection and quantification rates of SARS-CoV-2 RNA (N2-assay) analyzed from wastewater at 28 WWTPs in the groups of reported new COVID-19 case incidence rates in the corresponding sewer network area in Finland over a surveillance period of 10-months (3 August 2020 to 31 May 2021).

| Reported incidences (/100, 000) |  | Total samples included | N2 assay >LOD |  | N2 assay >LOQ |  |
| --- | --- | --- | --- | --- | --- | --- |
| Interval | Geometric mean cases |  | No of samples positive (n) | Positive rate (%) | No of samples positive (n) | Positive rate (%) |
| 0 | 0 | 39 | 6 | 15 | 1 | 3 |
| 0-2.50 | 1.6 | 20 | 6 | 30 | 0 | 0 |
| 2.5-3.64 | 3.01 | 20 | 5 | 25 | 0 | 0 |
| 3.64-4.76 | 4.39 | 20 | 6 | 30 | 1 | 5 |
| 4.76-6.67 | 5.53 | 20 | 10 | 50 | 0 | 0 |
| 6.67-8.90 | 7.96 | 20 | 8 | 40 | 0 | 0 |
| 8.90-11.64 | 10.23 | 20 | 14 | 70 | 1 | 5 |
| 11.64-14.29 | 13.05 | 20 | 16 | 80 | 0 | 0 |
| 14.29-16.39 | 15.23 | 20 | 14 | 70 | 0 | 0 |
| 16.39-17.50 | 17.07 | 20 | 13 | 65 | 0 | 0 |
| 17.50-20.47 | 18.91 | 20 | 16 | 80 | 0 | 0 |
| 20.47-22.95 | 21.63 | 20 | 17 | 85 | 2 | 10 |
| 22.95-27.17 | 25.46 | 20 | 17 | 85 | 0 | 0 |
| 27.17-30.16 | 28.78 | 20 | 19 | 95 | 2 | 10 |
| 30.16-32.79 | 31.65 | 20 | 17 | 85 | 0 | 0 |
| 32.79-38.10 | 35.34 | 20 | 18 | 90 | 2 | 10 |
| 38.10-42.42 | 40.83 | 20 | 19 | 95 | 1 | 5 |
| 42.42-47.27 | 45.09 | 20 | 19 | 95 | 3 | 15 |
| 47.27-52.38 | 49.84 | 20 | 19 | 95 | 3 | 15 |
| 52.38-57.14 | 54.60 | 20 | 17 | 85 | 6 | 30 |
| 57.14-62.86 | 59.72 | 20 | 19 | 95 | 4 | 20 |
| 62.86-74.50 | 69.67 | 20 | 20 | 100 | 5 | 25 |
| 74.50-80.58 | 77.46 | 20 | 20 | 100 | 4 | 20 |
| 80.58-94.49 | 88.12 | 20 | 20 | 100 | 7 | 35 |
| 94.49-100.00 | 96.79 | 20 | 20 | 100 | 7 | 35 |
| 100.00-110.81 | 106.82 | 20 | 20 | 100 | 9 | 45 |
| 110.81-121.21 | 116.36 | 20 | 19 | 95 | 11 | 55 |
| 121.21-137.00 | 126.47 | 20 | 20 | 100 | 13 | 65 |
| 137.00-163.67 | 149.23 | 20 | 20 | 100 | 11 | 55 |
| 163.67-196.50 | 177.96 | 20 | 20 | 100 | 15 | 75 |
| 196.50-238.72 | 220.67 | 20 | 20 | 100 | 14 | 70 |
| 238.72-299.19 | 271.32 | 20 | 20 | 100 | 15 | 75 |
| 299.19-460.70 | 356.81 | 20 | 20 | 100 | 18 | 90 |
| 460.70-767.50 | 521.05 | 14 | 14 | 100 | 10 | 71 |

Reported incidences = 14-day sum /100, 000 persons. >LOD = Samples with SARS-CoV-2 GC more than limit of detection with N2-assay. >LOQ = Samples with SARS-CoV-2 GC more than limit of quantification with N2-assay.

**Figure 2.**
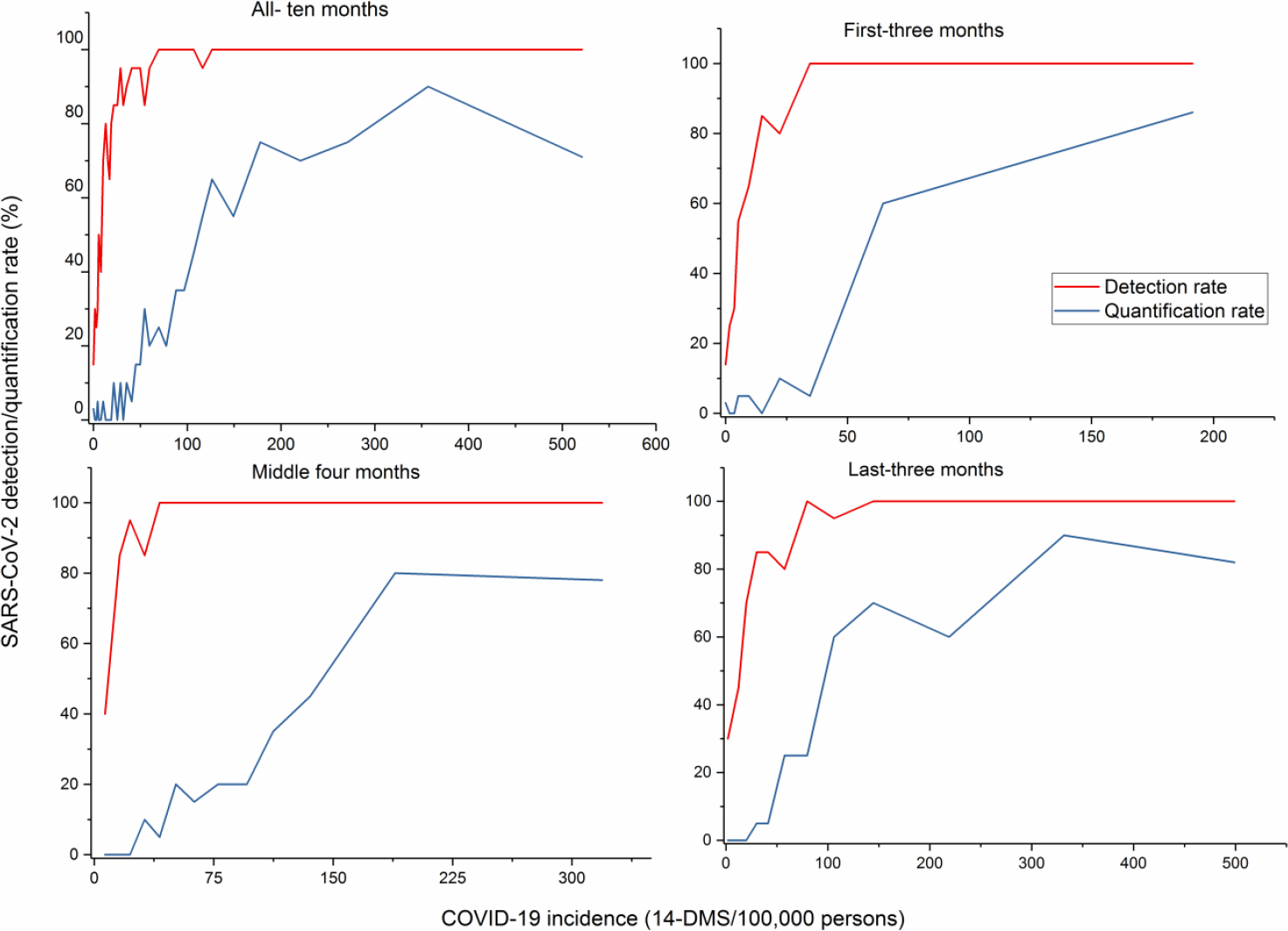
Relationship between COVID-19 incidence and the detection and quantification rates of SARS-CoV-2 RNA determined using N2-assay in wastewater.

**Figure 3.**
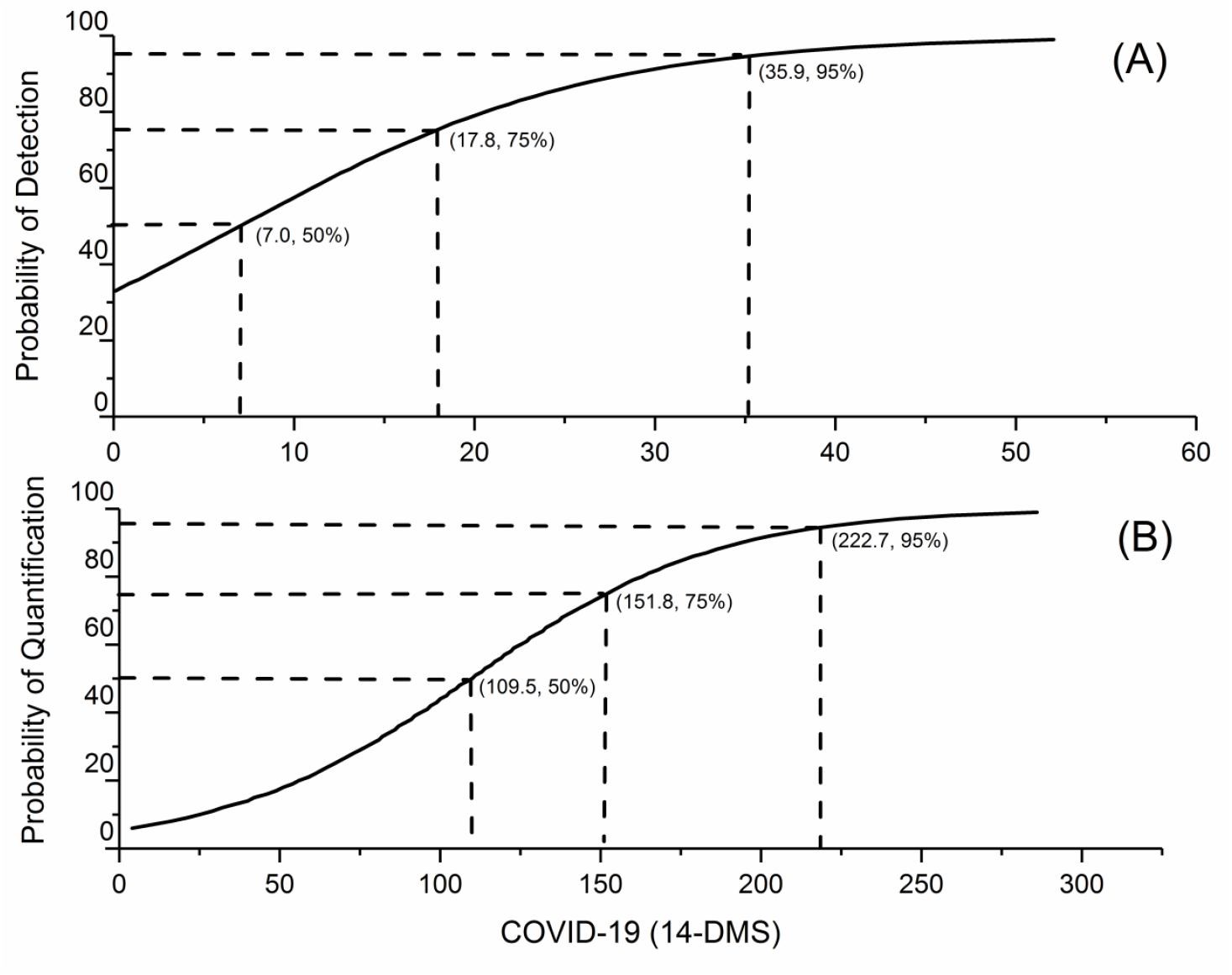
Fourteen-day COVID-19 incidence rate thresholds in the sewer network area covered by national surveillance in Finland required for wastewater-based SARS-CoV-2 detection and quantification estimated with logistic regression.

The quantification rate of SARS-CoV-2 RNA increased almost constantly as the COVID-19 incidence increased in the sewer network area but did not reach 100% during the study period (Table 2, Figure 2). In fact, as compared to the SARS-CoV-2 GC detection rate in WW, the quantification of SARS-CoV-2 required significantly more COVID-19 cases: the quantification rate was 50%, 75% and 95% when the 14-day incidence was 110, 152 and 223 COVID-19 cases, respectively, per 100 000 persons (Figure 3).

During the 10 months of the study period, a national view with WW data originating from 28 WWTPs is provided with ascending and descending phases of the COVID-19 incidence in the country. However, the incidence trend variations between the first three months (ascending phase), the middle four months (plateau phase) and the last three months (descending phase) displayed only minor changes in the WW-based detection and quantification rates (Figure 2).

During the study period, the COVID-19 incidence trend gradually increased from August 2020 when the mean (± SE) 14-DMS per 100 000 population was 4.5±0.7, until December when the 14-DMS was 102.9±15.7, decreased slightly in January 2021, (14-DMS was 79.4±6.6), increased gradually and reached the highest mean 14-DMS in March (172.0±19.4) and then decreased gradually by the end of the study period, May 2021, when 14-DMS was 63.2±8.0 per 100 000 population (Figure 1). Consistent with COVID-19 trends, the SARS-CoV-2 detection rate varied, being 38% in August and 91% in December, reaching its peak at 95% in February, and dropping to 90% in March and 71% in May. Similarly, the WW influent flowrate-normalized SARS-CoV-2 RNA copy number (mean ± SE) changed according to COVID-19 incidence trends: 7.00 ± 0.01 in August and 7.15 ± 0.15 in September, reaching its peak at 7.58 ± 0.07 in March and decreasing to the value of 7.36 ± 0.10 GC/person/day in May.

### 3.2 COVID-19 incidence determines the SARS-CoV-2 detection and quantification in wastewater at each WWTP

SARS-CoV-2 RNA was detected in 79% (including 6% uncertain) out of a total of 693 samples analyzed with N2 assay (Table 3). Among them, only 24% of the total samples had a GC above the LOQ, with a clear difference in the quantification rates between the large (33%) and small (12%) WWTPs.

**Table 3.**
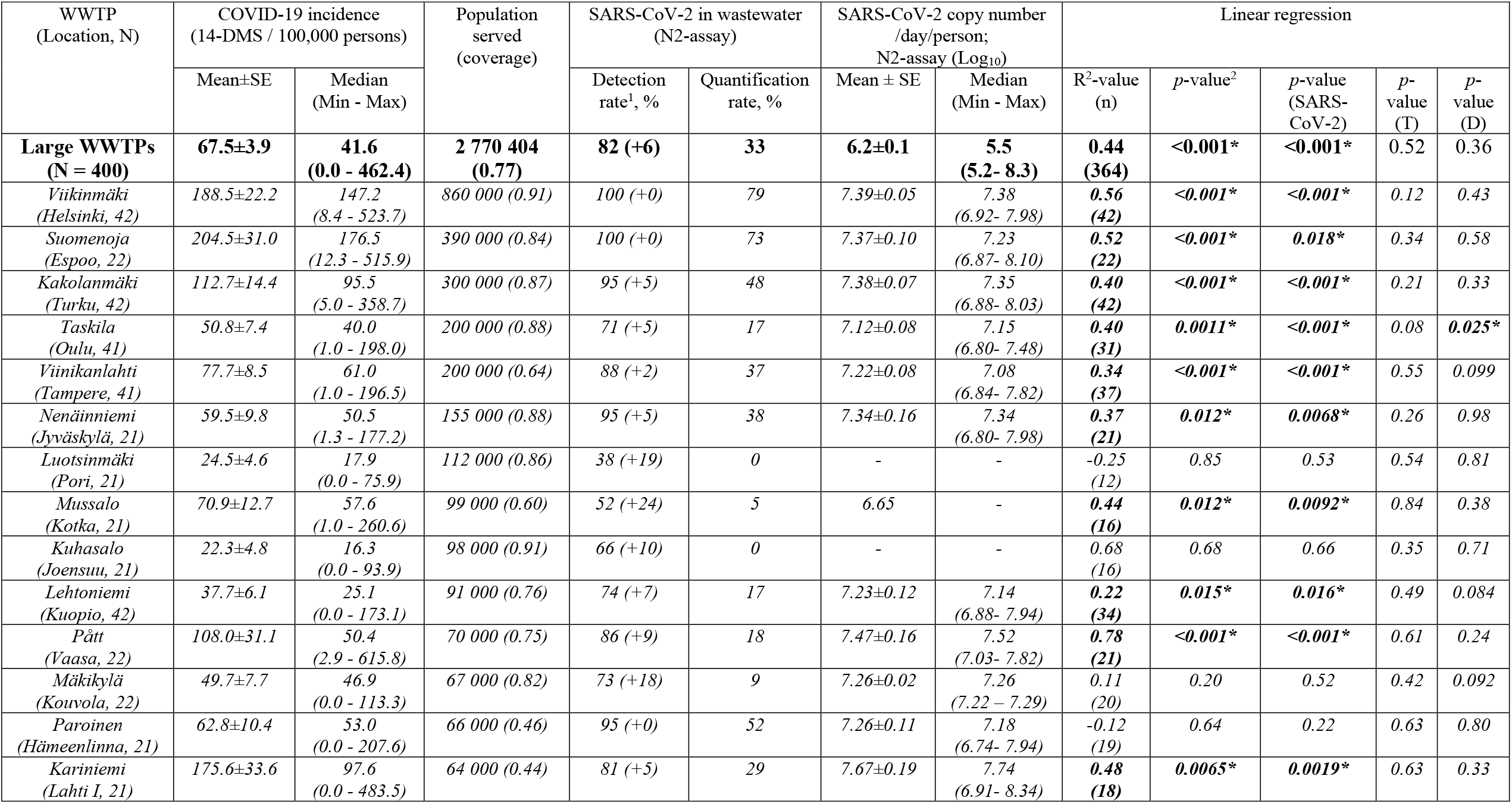
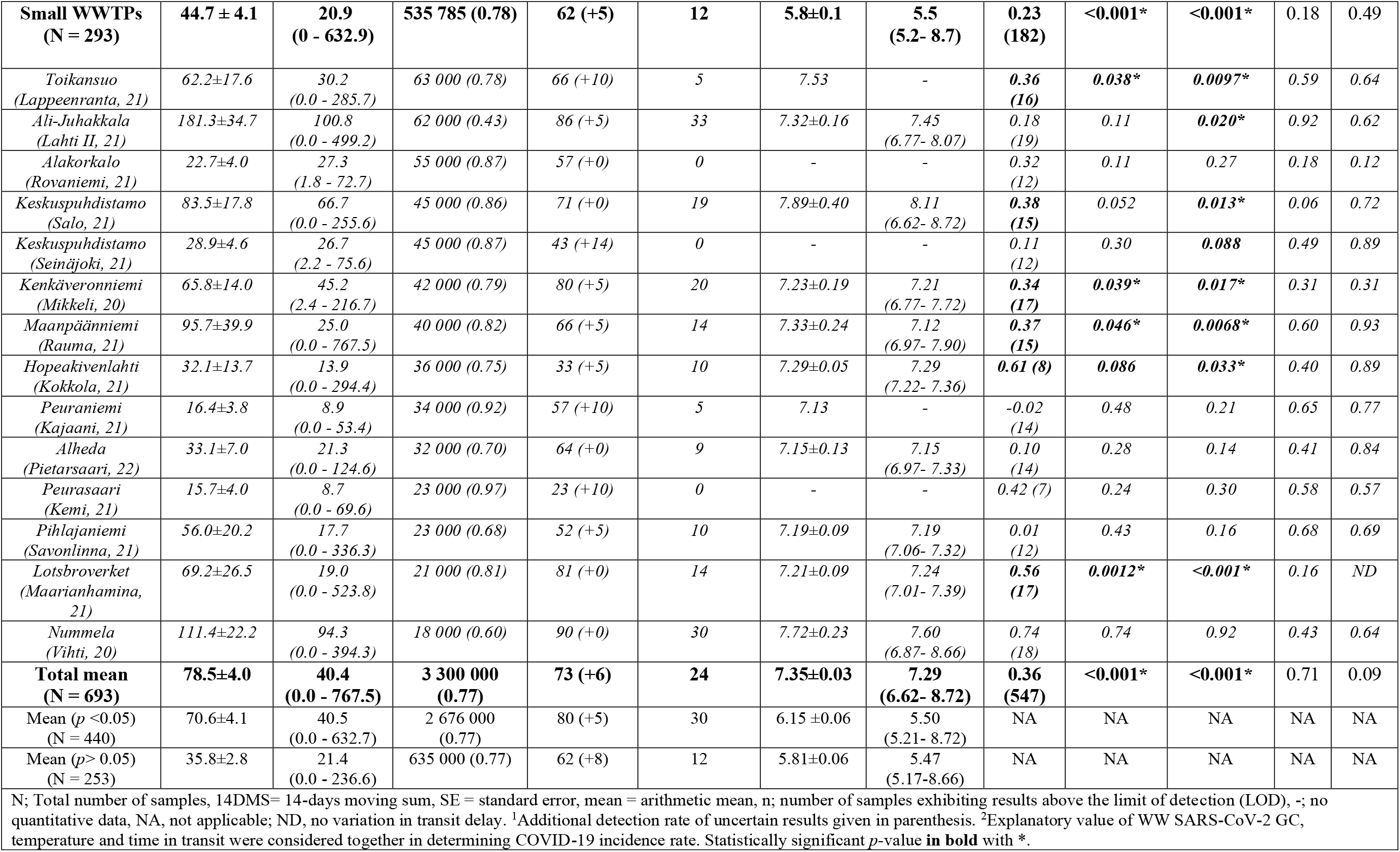
Reported COVID-19 cases in the sewer network areas, wastewater-based SARS-CoV-2 detection and quantification rates, SARS-CoV-2 gene copies enumerated with N2-assay, and linear regression analysis of 14-day moving sum as a dependent variable and flow-corrected SARS-CoV-2 gene copies/day/100 000 persons (SARS-CoV-2), sample temperature after transport (T), and sample processing delay (D) as explaining variables in A) large and B) small WWTPs, serving population above and below 63 500 inhabitants, respectively, out of the 28 WWTPs included in a 10-month national surveillance period in Finland.

The SARS-CoV-2 RNA detection rate varied in WWTPs located in different cities following the COVID-19 incidence (Table 3). The highest detection rates (95% or higher) of SARS-CoV-2 in WW were recorded in Helsinki, Espoo, and Turku, where the mean COVID-19 incidence rates per 100 000 persons exceeded 100, but also in Jyväskylä and Hämeenlinna, where the mean COVID-19 incidence was around 60. In all these locations, SARS-CoV-2 was quantifiable in at least 35% of the WW samples analyzed. However, cities with a small population, namely Vihti and Salo, exhibited the highest mean SARS-CoV-2 RNA GC counts in WW: 7.72±0.23 and 7.89±0.40 Log10 GC/day/person, respectively. At the other extreme, the WW-based SARS-CoV-2 GC never exceeded the LOQ and the median values of COVID-19 incidence were always less than 30 per 100 000 persons (14-DMS) in WWTPs of Pori, Joensuu, Rovaniemi, Seinäjoki, and Kemi, where the SARS-CoV-2 detection rates in WW were 57%, 76%, 71%, 57%, and 33%, respectively. At 13 WWTPs where the median of 14-day COVID-19 case incidence rate was 8.9–66.5 per 100 000 population, the SARS-CoV-2 RNA detection rate in WW was quite high, up to 95%, while the quantification rate remained less than 20%.

Linear regression was conducted to determine whether the number of customers or WW influent inflow volumes of WWTP, sample temperature after transport to the laboratory, sample collection month, or delay in sample processing can affect the relationship between the WW SARS-CoV-2 GC and the COVID-19 incidence. Of these, three factors; SARS-CoV-2 GC, sample temperature upon arrival at the laboratory, and sample processing delay collectively accounted for a 36% variation in overall COVID-19 incidence (Table 3). Sample temperature upon arrival at laboratory and transportation delay affected the *p*-value of the linear regression but SARS-CoV-2 GC was the only significant factor determining COVID-19 incidence alone.

The regression model indicated a significant relationship between the WW-based SARS-CoV-2 GC and the COVID-19 incidence in total at 15 out of 28 WWTPs (Table 3). At four out of the thirteen WWTPs where a significant relationship was not found, the GC of SARS-CoV-2 RNA remained below the quantification limit during the whole study period. In the other five WWTPs, the sewer coverage was less than 80% of the total population in the area and thus the COVID-19 cases may have been inhabitants from the areas not covered. In general, larger cities had higher mean COVID-19 incidence rates, and SARS-CoV-2 detection and quantification rates and mean copy numbers in wastewater (*p <* 0.001) and thus the linear regression were in better agreement in the group of large cities than small cities (Table 3).

Trends in reported new COVID-19 cases varied in the sewer network areas of the 28 WWTPs (Figures 4A-4G). The national capital region (Helsinki and Espoo) and the third-largest city and maritime gateway from Sweden, Turku, were constantly the major hotspots for COVID-19. These hotspots and other neighboring cities (Hämeenlinna, Lahti, and Vihti) had two major waves of COVID-19 during the surveillance period; the first wave in November–December 2020 and the main wave in March–April 2021 (Figure 4A and 4B). In other locations, namely the cities of Salo, Rauma, Maarianhamina, Kokkola, Savonlinna and Lappeenranta, there was a one-time COVID-19 peak in March–April 2021 (Figure 4C-4F). The north-western coastal cities of Vaasa, Pietarsaari, Rovaniemi, and Kemi had a one-time peak in October–November 2020 (Figure 4C, 4E, and 4G).

**Figure 4.**
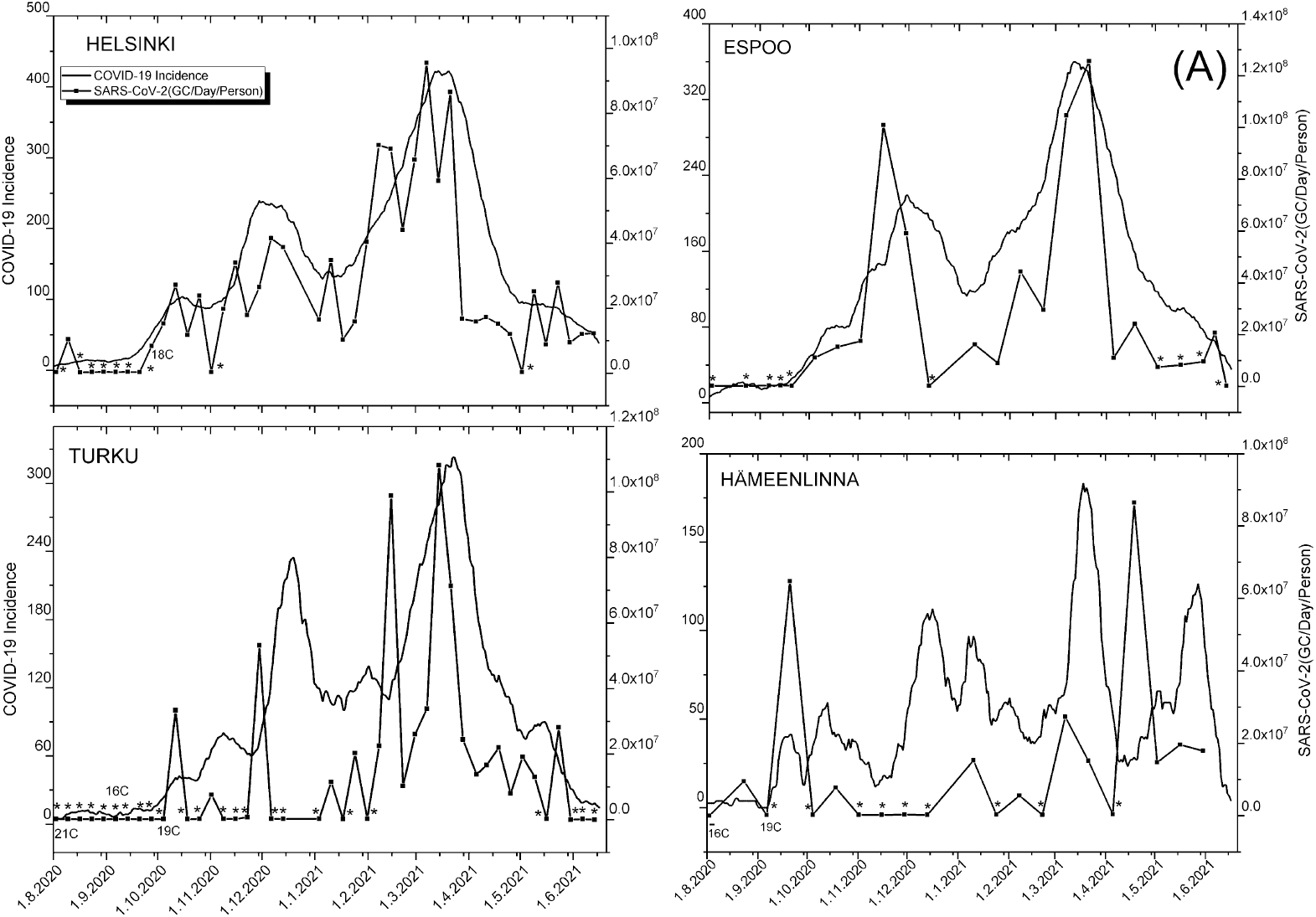
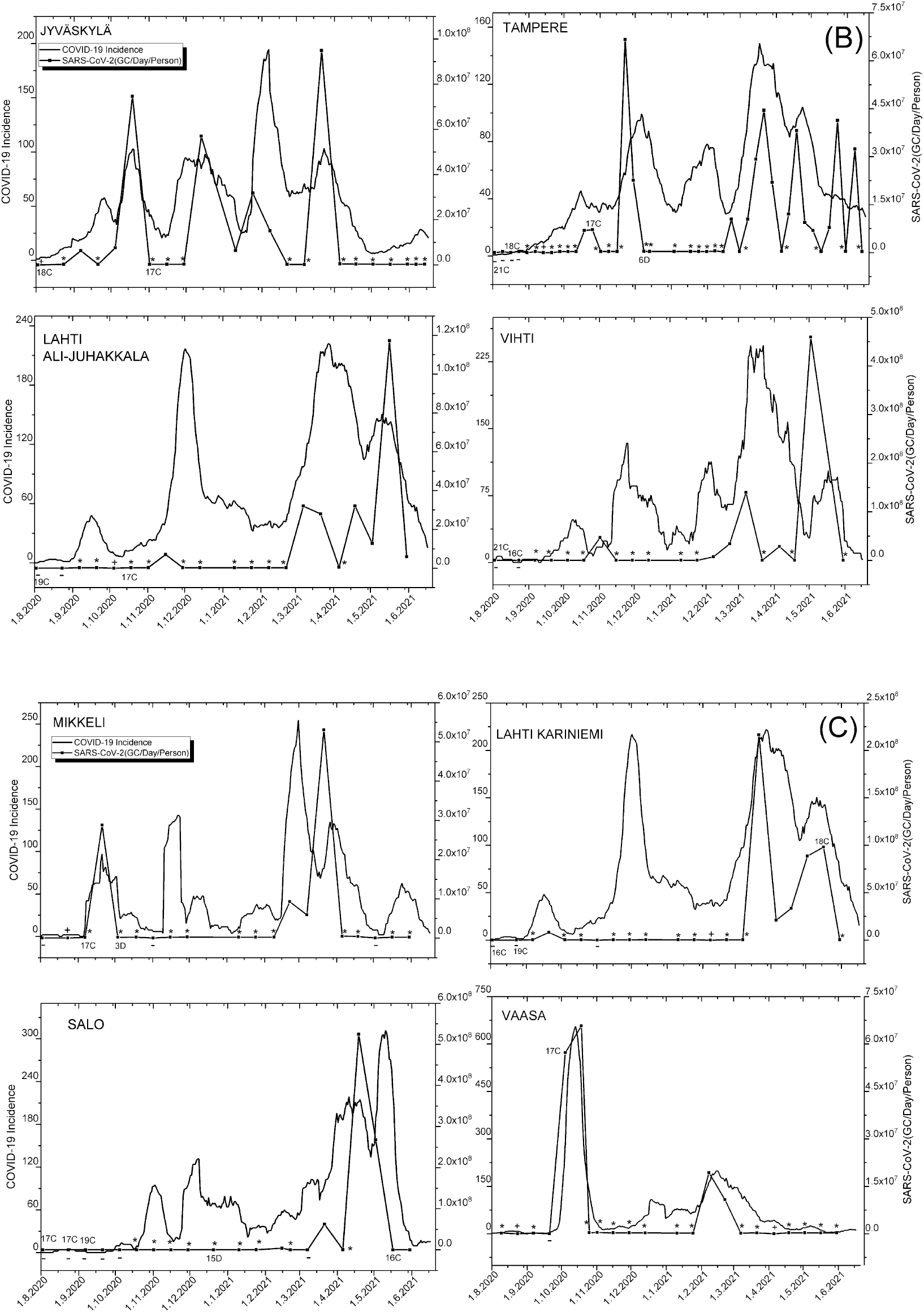
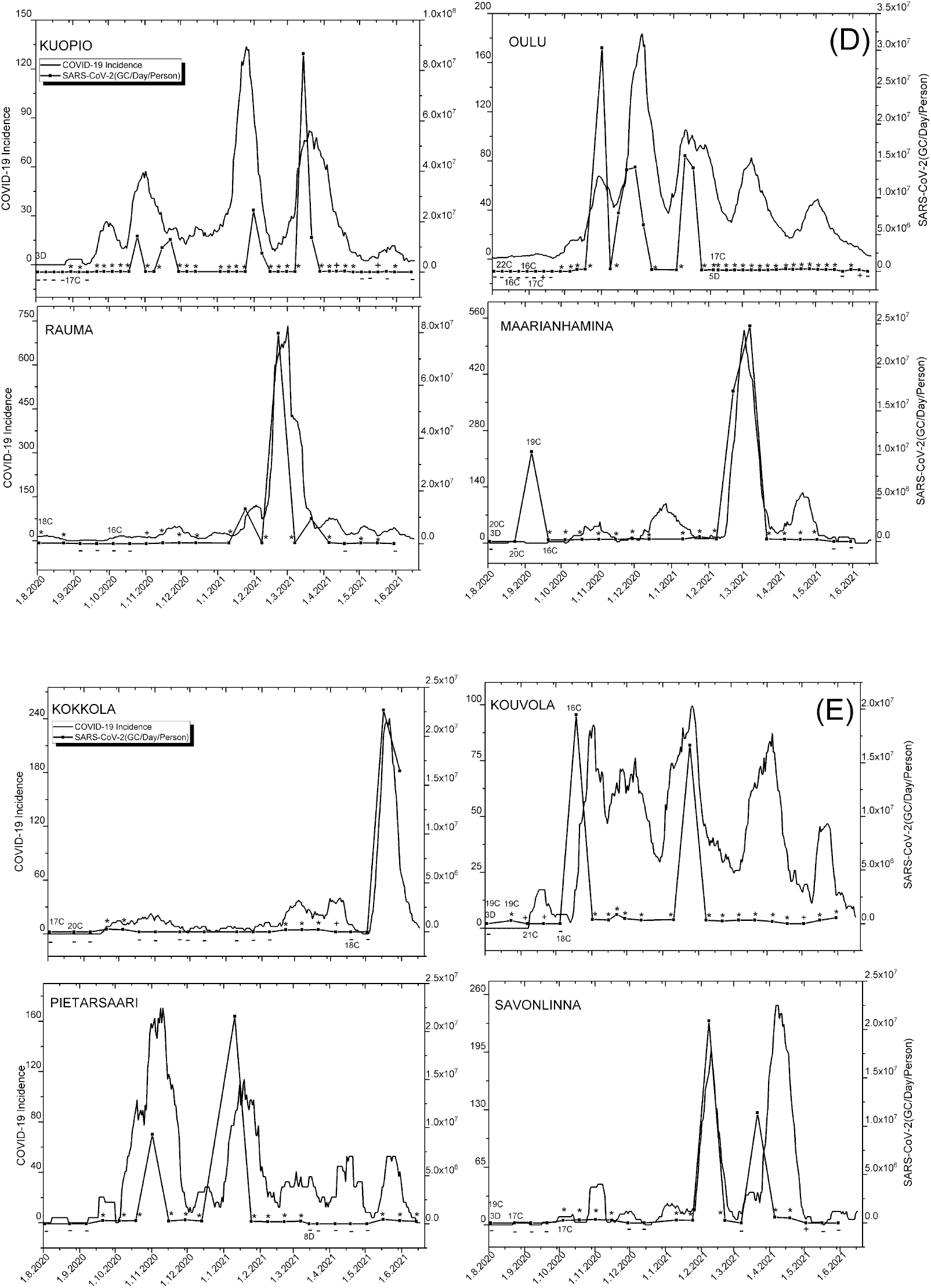
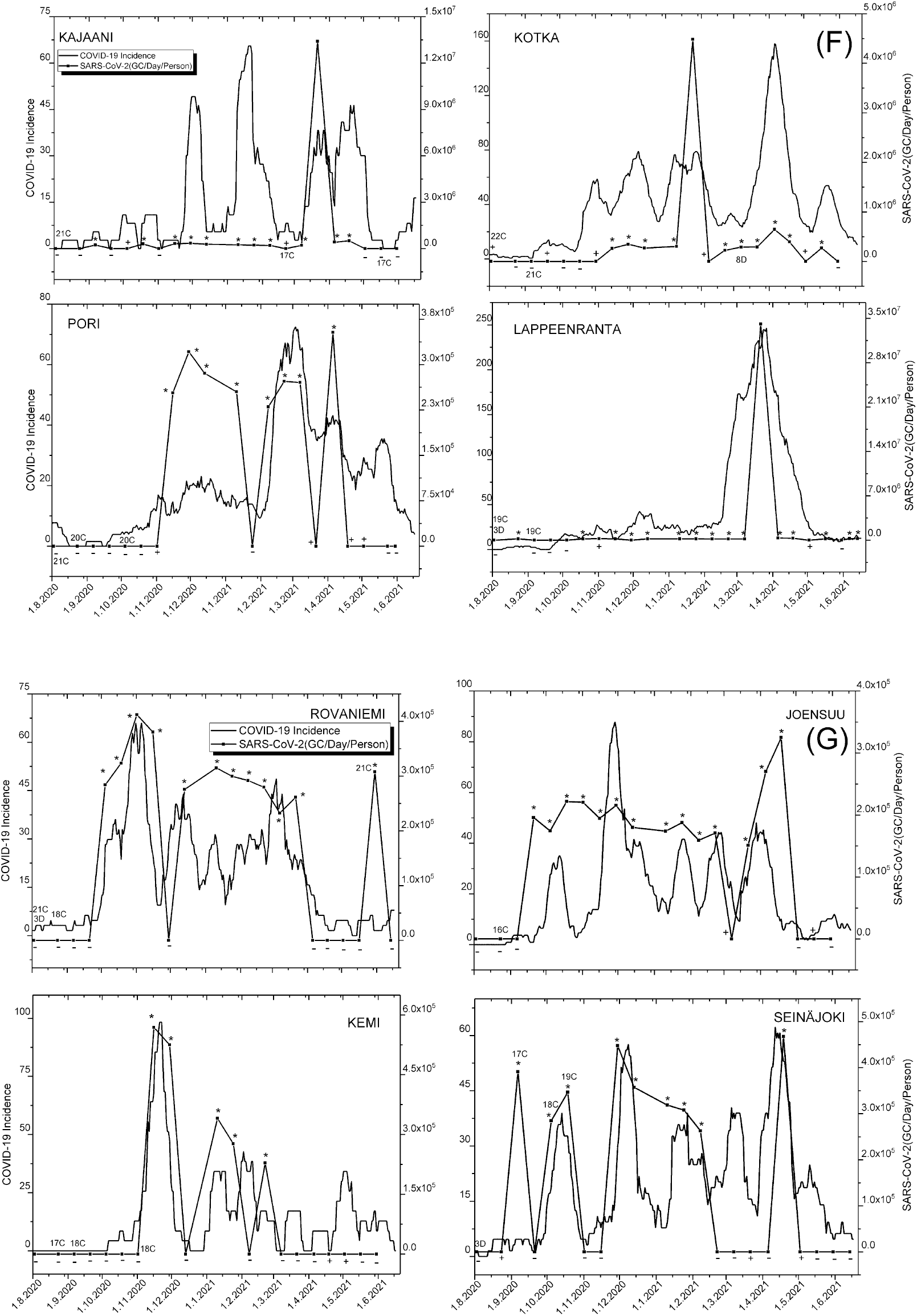
The trend of SARS-CoV-2 RNA (GC/day/person) in WW samples and COVID-19 incidence (per 100 000 persons). Sample dots with (-) denote SARS-CoV-2 was not detected, (+) denotes that the detection was uncertain, (*) denotes detected, but below quantification limit (LOQ). **A)** City areas (Helsinki, Espoo, Turku, Hämeenlinna) with quantification rate (QR) 48–79%. **B)** City areas (Jyväskylä, Tampere, Lahti II, Vihti) with QR 30–38%. **C)** City areas (Mikkeli, Lahti I, Salo, Vaasa) with QR 18–29%. **D)** City areas (Kuopio, Oulu, Rauma, Maarianhamina) with QR 14–17%. **E)** City areas (Kokkola, Kouvola, Pietarsaari, Savonlinna) with QR 9–10%. **F)** City areas (Kajaani, Kotka, Pori, Lappeenranta) with QR 0–5%. **G)** City areas (Rovaniemi, Joensuu, Kemi, Seinäjoki) with QR 0%. Caution: the Y-axis of each graph is different, and the x-axis is the same. Samples with arrival temperature higher than 15 °C (C) and transportation delay more than or equal to three days (D) are shown.

### 3.3 SARS-CoV-2 viability and methodological aspects in wastewater

The cell culturing attempts for the selected WW samples indicated that the SARS-CoV-2 target was non-viable in the samples analyzed.

Further, the WW-based SARS-CoV-2 RNA assay target was found to significantly affect the surveillance outcomes. Out of the total of 386 samples analyzed using both N2 and E-Sarbeco assays, the detection rate with N2 assay was significantly higher than with E-Sarbeco assay [*Χ*^2^ (1)> = 183.4, *p* < 0.0001]. All except four samples that were positive for SARS-CoV-2 with E-Sarbeco assay were also positive with N2 assay. Notably, 14% of samples proven to contain SARS-CoV-2 with N2 assay were assigned as false-negative with E-Sarbeco assay. Even though Spearman’s correlation coefficient (ρ) between the detection of SARS-CoV-2 RNA with N2-assay and E-Sarbeco assays was 0.676 (*p* < 0.0001), SARS-CoV-2 RNA was consistently more frequently detected with N2-assay than with E-Sarbeco assay throughout all sampling months. Due to the lower sensitivity of the E-Sarbeco assay, it was no longer used after January 2021 for the national WW-based surveillance.

The multiple linear regression analysis conducted indicated that crAssphage is an insufficient normalization method for WBE since a significant relationship between 14-day COVID-19 case incidence rate and crAssphage corrected SARS-CoV-2 GC numbers in WW was found only for two out 28 WWTPs. The corresponding relationship when using flow-corrected SARS-CoV-2 numbers was found in total at 15 out of 28 WWTPs (Table 3). However, crAssphage assay seemed to provide useful results in the ultrafiltration method performance testing, where this DNA-based assay resulted in higher recovery rates Ultrafiltration recovery efficiencies (68.9-110.3%) as compared to the mengovirus, a non-enveloped RNA-virus, recovery (0-76.9%).

## 4. Discussion

This study reports the surveillance results of SARS-CoV-2 RNA analyzed from 28 WWTPs in Finland between August 2020 to May 2021. By using cell culture-based viability assays for selected samples, the study also provides further evidence that SARS-COV-2 coronaviruses do not pose a waterborne transmission risk as they remain non-viable in community wastewater influents as stated by others (Rimoldi et al. 2020, Westhaus et al. 2021, Ahmed et al. 2021c).

By analyzing a total of 693 WW samples, we found that WW-based detection of SARS-CoV-2 from multiple locations is an effective measure for tracing national trends in COVID-19. By using the ultrafiltration method and N2 RT-qPCR assay, the relative quantification of SARS-CoV-2 numbers was possible at 50% probability, when the COVID-19 incidence rate in the sewer network area exceeded 152 cases per 100 000 persons. The SARS-CoV-2 detection reached 50% probability already when the preceding 14-day COVID-19 case incidence rate was about seven cases per 100 000.

The COVID-19 trend was highly variable between the sewer network areas throughout the country, with the large cities (Helsinki, Espoo, and Turku) having the most influence on the national trends of COVID-19 incidence. These large cities were consistently the nation’s pandemic hotspots, which is not surprising as these cities are the major financial, tourist, and education hubs and gateways to the country from abroad. Overall, the detection trend of SARS-CoV-2 RNA in WW had a strong relationship with reported new COVID-19 cases in the sewer network area and showed good agreement with earlier studies (Wang et al. 2020, Gonzalez et al. 2020, Westhaus et al. 2021). The monitoring of SARS-CoV-2 RNA in WW captured both trends and peaks of COVID-19 incidences, both at local and national levels. The WW-based SARS-CoV-2 surveillance data often followed the same sequence: SARS-CoV-2 was first detected as an uncertain observation, then detected but below LOQ quantities, and finally quantified after a further increase in COVID-19 incidence, illustrating the simultaneous spread of COVID-19 cases in individual testing among the inhabitants of the studied sewer network areas.

Although traces of SARS-CoV-2 in WW might remain below the limit of detection (LOD) or at least below the limit of quantification (LOQ) of the method (Ahmed et al. 2020, Ahmed et al. 2021b), the data presented herein demonstrates a clear connection between the change in COVID-19 incidence noted in the individual testing and the presence and quantity of SARS-CoV-2 RNA in WW. In communities where COVID-19 infection rates are low, RNA is less likely to be quantified in WW and the reporting systems need to function with the available binary results (SARS-CoV-2 detected/non-detected). In our study, the sole reliance on the quantified SARS-CoV-2 GC WW results would have caused a loss of most of our results (∼76%). In the WWTPs of five cities (Pori, Joensuu, Rovaniemi, Seinäjoki, and Kemi, where 14-day COVID-19 case incidence rate was always less than 27), SARS-CoV-2 RNA was not quantified in any of the studied WW samples. Indeed, some earlier studies have also reported a poor correlation between COVID-19 incidence with SARS-COV-2 RNA copies mainly during periods of low COVID-19 incidence (Hillary et al. 2020, Ahmed et al. 2021a).

In our study, only six out of 693 WW samples were positive with SARS-COV-2 RNA when the reported 14-day COVID-19 case incidence was zero. While the detection of SARS-CoV-2 RNA in WW influent infers there being at least one person with COVID-19 shedding SARS-CoV-2 into the WW in the sewer network area, the NIDR collects the individual test positivity results based on the municipality of residence. However, due to the pandemic and the recommendation not to travel, the movement of people between municipalities could have been during the study period lower than usual. Further, although individual COVID-19 tests were available for all symptomatic people in Finland over the study duration, not everyone is willing to be tested and the infected person can also be asymptomatic, pre-symptomatic, or post-symptomatic.

Conversely, the non-detection of SARS-CoV-2 RNA in WW influents does not guarantee the absence of infected people in the sewer network area. The possible reasons for non-detection in WW can be due to (a) an absence of infected people, (b) the virus load is below the WW method LOD, or (c) the periodic flow of the virus with limited numbers of infected people has not been captured during the period of sample collection. Further, the mixing with other WW flows, stormwater infiltration, the diurnal variation in shedding, and hydraulic residence time in the sewer collection system, can also affect the probability of detection of SARS-CoV-2 RNA, particularly in low prevalence conditions (Ahmed et al. 2020, Hillary et al. 2021). The higher sensitivity of N2 assay than E-Sarbeco assay in our hands and some contradictory results reported earlier in relation to N2 assay performance (Ahmed et al. 2021a; Gerrity et al. 2021) highlights the need for special attention during the selection of RT-qPCR assay for SARS-CoV-2 determinations from WW.

## 5. Conclusions

- This study shows a clear relationship between the 14-day COVID-19 case incidence rate in sewer network areas and the detection and quantification rate of SARS-CoV-2 RNA in WW influent samples of respective WWTPs.
- The 14-day COVID-19 case incidence of 7.0 per 100 000 persons yielded about 50% probability of detecting SARS-CoV-2 from wastewater samples, and a 95% wastewater detection rate was reached when the COVID-19 incidence was about 36 cases per 100 000 persons.
- A much higher 14-day COVID-19 case incidence rate was required to quantify than to detect SARS-CoV-2 from the wastewater samples. 50% and 95% probability to quantify SARS-CoV-2 in wastewater was achieved when the 14-day COVID-19 case incidence was 110 and 223 per 100 000 persons, respectively.
- This finding supports the use of binary (detected/not-detected) results of SARS-CoV-2 RNA monitoring as a basis of WW-based surveillance results reporting during periods of low COVID-19 incidence.
- This study did not find viable SARS-CoV-2 particles in WW samples.

## Data Availability

This work is part of that carried out by the national COVID-19 task force at the Finnish Institute for Health and Welfare (THL). The wastewater-based surveillance results of SARS-CoV-2 in comparison to detected COVID-19 cases in the sewer network areas and the wastewater data is publicly available [In Finnish]. Further, the numbers of confirmed new COVID-19 cases reported in each municipality of Finland are publicly available.

https://www.thl.fi/episeuranta/jatevesi/jatevesiseuranta_viikkoraportti.html

https://thl.fi/en/web/thlfi-en/research-and-development/research-and-projects/sars-cov-2-at-wastewater-treatment-plants/coronavirus-wastewater-monitoring

https://thl.fi/coronamap

https://thl.fi/en/web/infectious-diseases-and-vaccinations/what-s-new/coronavirus-covid-19-latest-updates/situation-update-on-coronavirus/map-application-on-corona-cases

## Author contributions

AT, AL and AH have contributed equally on the work. AT wrote the first draft of the manuscript; AL conducted the initial RT-qPCR result calculation and data interpretation; and AH was in charge of the wastewater collection and analysis. AL, AH, AS, and AR undertook the wastewater investigations, processing, and analysis of the laboratory data. AL, AH, OL, and AJ undertook the processing and analysis of the epidemiological data. PO and HA were in charge of the cell culture work in SARS-CoV-2 viability assessments. IM and CSK provided resources for the team. AT, AL, OL, and TP led the data interpretation and the writing of the manuscript. TP and CSK conceptualized the project, supervised the work, and oversaw funding acquisition. TP prepared and all authors contributed to the final draft of the article.

## Acknowledgements

The authors would like to express special thanks to Marjo Tiittanen, Mirka Korhonen, Mari Turunen, Tarja Rahkonen, Tiina Heiskanen, Eveliina Nurmi, Kristiina Valkama and Arja Moilanen for their assistance in the laboratory, and Aino Kankaanpää for her help with sample transportation arrangements. The personnel of the wastewater treatment facilities in all 28 wastewater-based SARS-CoV-2 surveillance locations in Finland are highly acknowledged for their support and timely efforts for wastewater composite sampling and sample transportation arrangements for the work. Further, we acknowledge the NORMAN Network, COVID-19 WBE Collaborative, and European Commission Initiative “SARS-CoV-2 monitoring employing sewers” for sharing their knowledge and resources for this work.

